# Sleep disturbances and related factors among preschool children in rural areas of China: a cross-sectional study

**DOI:** 10.1101/2020.10.20.20215806

**Authors:** Tianming Zhao, Kun Xuan, Haixia Liu, Xin Chen, Guangbo Qu, Yile Wu, Jian Zhang, Yehuan Sun

## Abstract

**Objective:** Sleep disturbances have been serious since they are believed to be associated with various health problems in preschool children. In this study, we aimed to explore the prevalence of sleep disturbances amongst preschool children in rural areas of China and examine the underlying associated factors.

**Methods:** A cross-sectional study was conducted in rural areas of Anhui province, China from September 2019 to January 2020. To explore the potential associated factors comprehensively, caregivers of children completed a structured questionnaire, the Children’s Sleep Habits Questionnaire (CSHQ), the Strengths and Difficulties Questionnaire (SDQ), Self-rating Anxiety Scale (SAS) and the Chinese version of the adapted Identification and Management of Feeding Difficulties (IMFeD). One-way ANOVA and hierarchical multiple regression are conducted to explore the potential influencing factors of sleep disturbances in preschool children.

**Results:** About 89.3% of the preschool children aged from 3 to 6 years had potential sleep disturbances (scored above the cutoff value). Besides, ages of children, discipline attitudes of father and mother, main educational methods, caregivers of children, caregiver anxiety and hyperactivity/inattention, prosocial behavior, conduct problems, emotional symptoms, peer problems, food preference, fear of feeding, parental misperception and organic disease all contributed significantly to CSHQ total score by accounting for approximately 27.4% (R^2^=0.274) of the variation.

**Conclusions:** Our study indicated that the prevalence of sleep disturbances among preschool children in rural area of China is quite high. Furthermore, the potential risk factors are complicated which include factors related to both children and caregivers especially their parents.

## Introduction

Sleep disturbances are associated with various cognitive, behavioral, and emotional difficulties in children and the link between them are complex [1-3]. Preschool age is a sensitive developmental stage and obtaining adequate amounts of sleep is vital to their social, emotional, behavioral, and academic development [4]. In this period, improper parental supervision such as inadequate care, parental absence and overprotection may have long-term effects on their children’s development and health outcomes such as sleep disturbance even in their adulthood [5, 6]. Nevertheless, the characteristics of sleep practice, sleep patterns and sleep disturbances in different countries varied greatly especially between West and East [7]. Studies indicated that parents were more likely to report sleep problems in their children in Asian countries (24%) compared to their counterparts in Western countries (18%), and in Asian countries, parents’ perception in their children’s experience of sleep problems ranged from 15.1% in Korea to 43.7% in China [8]. A cross-cultural study between Chinese and Japanese preschool children has found difference in sleep patterns between two countries, although there was no difference in total severity of sleep disturbance between two countries, Chinese preschoolers were more severe in nighttime awakenings and sleep disordered breathing (SDB) whereas Japanese preschoolers were more susceptible in bedtime resistance [9]. Given the fact that the reason of prevalent discrepancy in different countries are still not clear enough, more detailed, and culture-tailored research is needed to implement in different countries and regions with different sociocultural backgrounds [10].

It is commonly believed that sociocultural factors including political conditions, economic considerations, ideological beliefs regarding societal and parental awareness may play an important role in sleep disturbances among preschool children [7, 10, 11], indicating that parents play an important role in ensuring the quality of their children’s sleep. Previous study has proven that parenting behavior is associated with children’s sleep duration [12]. In China, children tend to sleep shorter and wake up earlier than their counterparts in the USA; the potential reason may be the difference in school schedules, homework load and sleep habits of the 2 countries [13]. Therefore, the sleep problems of preschool children need to be paid more attention to.

In our study, we conducted a cross-sectional study aiming to explore the sleep disturbances prevalence and comprehensively determine the associated factors amongst preschool children in rural area of China based on a larger sample in Anhui province using several broadly validated and structured instruments. Our study will illuminate the distribution of sleep disturbances and related factors in preschool children in rural areas of China and supplement the previous study results.

## Methods

### Study design and subjects

We conducted this cross-sectional study from September 2019 to January 2020 and four counties (Funan county of Fuyang city, Changfeng county and Fexi county of Hefei city, Bowang of Maanshan city) were selected as our research sites based on the geographical locations of Northern, Central and Southern Anhui province. Preschool children from 26 kindergartens were enrolled in our study and their primary caregivers or their parents were asked to complete a structured questionnaire through face-to-face interview or filling by themselves based on their education level. In our study, preschool children were defined as children between the ages of 3 and 6 who were not yet attending primary school. Besides, we excluded the participants whose caregivers cannot communicate, such as cognitive impairment and deafness, and children with severe physical and mental illness were also excluded. After completing the questionnaire, all the items were checked to ensure their integrity and logic. A total of 3,802 questionnaires were distributed in our investigation, after checking for completeness and logic, 166 invalid questionnaires were excluded and 3,636 questionnaires were eventually included with the response rate of 95.6%. This study was approved by Ethics Committee of Anhui Medical University and followed the principles of informed consent and strict confidentiality.

### Assessment instruments

In this study, our questionnaire is well-established which consists of several scales to comprehensively evaluate the sleep problems among preschool children. Details of scales included in our questionnaire are as follows.

### Demographic questionnaire

We evaluated the sociodemographic information of children and their family by using this questionnaire which included such content: the general characteristics of the child (name, gender, date of birth, current weight, current length, daily screen time, etc.), the basic characteristics of the parents (age, height, weight, education, occupation, etc.), the basic situation of the family (family structure, sibling status, family income status, etc.) In our study, the definition of screen time level was set according to guidelines of the American Academy of Pediatrics (AAP) [14, 15]: normal screen time (≤60 min/d), excessive screen time (60 min/d < screen time ≤ 120 min/d) and severely excessive screen time (>120 min/d). According to the WHO standard of BMI for preschoolers [16, 17], for same gender and age, BMI between 85th percentile and 95th percentile was overweight, and greater than 95th percentile was obesity. Other information was self-reported by the respondent.

### The Children’s Sleep Habits Questionnaire (CSHQ)

In this study, the Children’s Sleep Habits Questionnaire (CSHQ) was used to assess the sleep problems of preschool children. The Children’s Sleep Habits Questionnaire (CSHQ) was developed in the USA by Owens et al which is widely utilized to assess and detect the sleep problems among children aged 4-10 years [18, 19]. In this study, we utilized Chinese version of CSHQ [20], which included 33 items describing sleep disturbance and items are rated through 3-point scale: “usually” if the sleep behavior occurred 5 to 7 times per week; “sometimes” for 2 to 4 times per week; and “rarely” for 0 to 1 time per week. Among those 33 items, the sleep disturbances are evaluated on eight subscales including bedtime resistance, sleep onset delay, sleep duration, sleep anxiety, night wakings, parasomnias, sleep-disordered breathing, and daytime sleepiness, and higher total and subscale scores represent more severe disturbances [7]. Parents or other caregivers are asked to recall the sleep behaviors of included child occurring over a “typical” recent week [21]. Given the fact that CSHQ is not regarded as a diagnostic questionnaire but as a screening tool for existing sleep disturbances, we utilized the definition of previous studies which defined total score > 41 as the potential clinical sleep disturbances [7, 21]. And the cutoff value for subscales are defined as scores >2 standard deviations above the published community control reference mean values [18] (bedtime resistance >10.84, sleep onset delay >2.31, sleep duration >5.27, sleep anxiety >7.79, night wakings >5.29, parasomnias >10.61, sleep-disordered breathing >4.50, daytime sleepiness >15.24) according to previous studies [10, 22, 23], Cronbach’s α coefficient was 0.668 for the full scale in this study.

### The Strength and Difficulty Questionnaire (SDQ)

The Strength and Difficulty Questionnaire (SDQ) was designed by Goodman and colleagues to evaluate behaviors, emotions and hyperactivity situations of children aged from 4 to 16 years [24]. And Chinese version of parental SDQ [25] was adopted to assess children’s emotional and behavioral question in our study. There are 25 items in this scale, which consists of four difficulty subscales (Emotional Symptoms, Conduct Problems, Hyperactivity /Inattention and Peer Relationship Problems) as well as a strength subscale (Prosocial Behavior). Each item has a score of 0∼2, with a rating of 3 levels: 0 for not match, 1 for a little match, and 2 for a complete match. Total difficulties score is the sum of the scores of the four difficulty subscales, and higher scores implying more difficulties; conversely, higher scores in Prosocial Behavior indicate more strength [10]. Cronbach’s α coefficient was 0.689 in this study.

### Self-rating Anxiety Scale (SAS)

In our study, we utilized Self-rating Anxiety Scale to evaluate the anxious status of caregivers. Self-rating Anxiety Scale is a self-report scale included 20 items covering a variety of anxiety symptoms [26]. Respondents are asked to score each of the items regarding to how it applied to them during the past week (1. None or A little of the time; 2. Some of the time; 3. Good part of the time; 4. Most or All of the time); higher score indicated more severe anxious status [27]. The scores of 20 items are calculated to obtain a raw score ranging from 20 to 80, and the standard score is generated through the raw score multiplied by 1.25; the standard score ≧50 indicates the presence of anxiety status; the standard score 50-59, 60-69 and ≧70 are considered mild, moderate and severe anxiety, respectively [28, 29]. In this study, the Cronbach’s α coefficient was 0.558.

### Identification and Management of Feeding Difficulties (IMFeD)

The Chinese version of adapted IMFeD tool was used to explore the feeding difficulties among preschool children [30, 31]. This questionnaire consists of 17 items which describe the feeding difficulties status of children through six dimensions: poor appetite (4 items), food preference (3 items), poor eating habit (4 items), fear of feeding (2 items), parental misperception (2 items) and organic disease (2 items). Each item is rated through 5-point scale (Behaviors happened every day; Behaviors happened 5-6 days per week; Behaviors happened 3-4 days per week; Behaviors happened 1-2 days per week; Behaviors never happened). Each item represents a problem in eating behaviors, and caregivers are asked to report how often their children had displayed eating disorder behaviors in each week according to each item; if any item is reported to occur always or often (Behaviors exceed 3 days a week), we considered the child had a problem in that dimension [31]. In this study, the Cronbach’s α coefficient was 0.887 for the full scale.

### Statistical analysis

The data entry was carried out by EpiData 3.2. SPSS 23.0 software were utilized to perform statistical analysis. Descriptive statistics were used to illustrate the distribution of participants in different sociodemographic characteristics through mean, standard deviation, frequencies, and percentages. One-way ANOVA and independent-samples t-test were used to compare the total score and score of each subscale of CSHQ among different levels of categorical sociodemographic factors. Pearson’s correlations were used to determine specific associations between subscales of CSHQ and subscales of SDQ as well as IMFeD. Furthermore, using total score of CSHQ as the dependent variable, hierarchical multiple regression was performed to explore the link between various independent variables and the dependent variable by entering independent variables successively: step 1 included sociodemographic factors; step 2 included anxious status of caregivers (evaluating by SAS); step 3 included subscales of SDQ; and subscales of IMFeD were included in step 4. Collinearity diagnosis was also performed to adjust the inter-correlation between independent variables. P<0.05 was considered as statistically significant.

## Results

### General characteristics and sleep disturbances of participants

In our study, the mean age of participants was 4.02 (SD=0.9) years with a range of 3-6 years. As shown in Table 1, among the participants approximately 54.2% were male and 45.8% were female. Regarding current left-behind status (left-behind children were defined as children who had been left behind by at least one parent for more than 6 months), about 36.2% were left-behind children. In birth weight, most of children (89.9%) in our study were in normal status (2500-4000g) as well as 4.3% underweight (<2500g) and 4.8% (>4000g) overweight children at birth. As for age, about 34.5% were 3 years old and 34.3% were 4 years old; 26.0% were 5 years old as well as 5.2% were 6 years old. In terms of BMI, most of children (83.3%) were in normal level with only 9.4% and 7.3% of them were overweight and obesity, respectively. More than half of father (55.4%) and mother (59.9%) attained an education level no greater than junior high school, with 44.4% of father and 39.8% of mother were high school or higher. When it comes to discipline attitudes of father and mother, nearly half of them (48.2%) were unanimous or occasionally inconsistent (47.0%), with only 4.9% of them were often inconsistent. In main educational methods, persuade education accounted for 82.9%, with 5.2% for natural education and 11.9% for scolding education. In terms of caregiver anxiety, most of caregivers (90.2%) reported no anxiety according to the result, with 7.1%, 1,8% and 0.3% for mild anxiety, moderate anxiety, and severe anxiety respectively. Most of caregivers (74.8%) in our study were parents, and 23.6% of them were grandparents as well as 1.6% of them were other relatives. 31.5% of participants were only child in their families, and about 68.5% of them had siblings. With regard to family average income monthly (CNY), approximately 38.0% of them were 5,000–10,000 and 32.5% were 3,000–4,999; 15.7% were >10,000 as well as 11.8% were under 3,000. As for screen time level, nearly half of children (46.1%) were in excessive screen time (EST) status (60 < ST≤120min), with about 25.4% of them were in normal levels (ST≤60 min) and 28.5% were in seriously excessive screen time (SEST) status (ST>120 min). Among preschool children in our study, 49.1% were picky eaters and 50.9% were not. Total scores of CSHQ in different groups of age, discipline attitudes of father and mother, main educational methods, guardian anxiety, screen time level and picky habits showed significant difference according to ANOVA analysis and t-test. Details of subscales of CSHQ are given in Table 1.

**Table 1.**
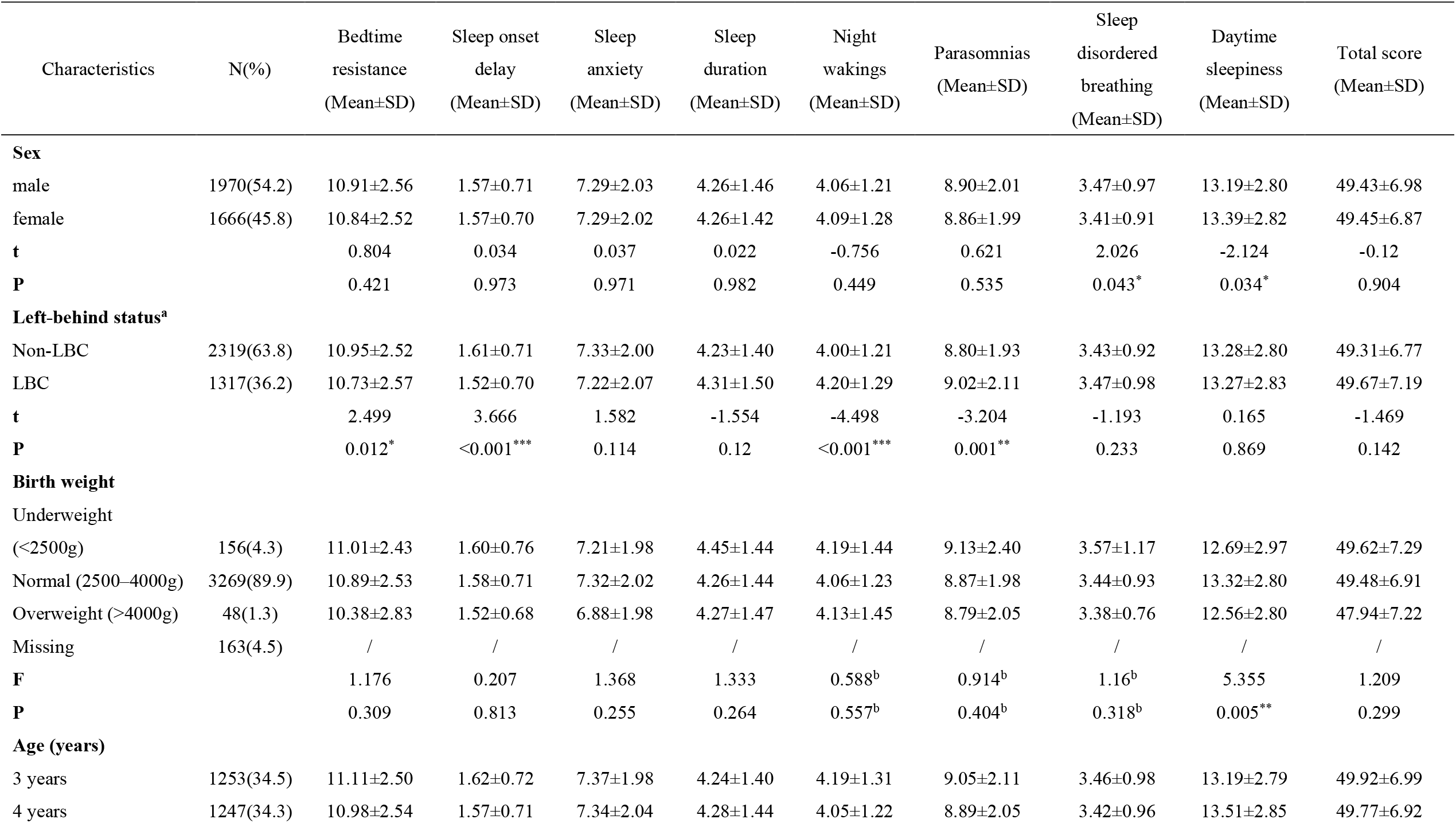
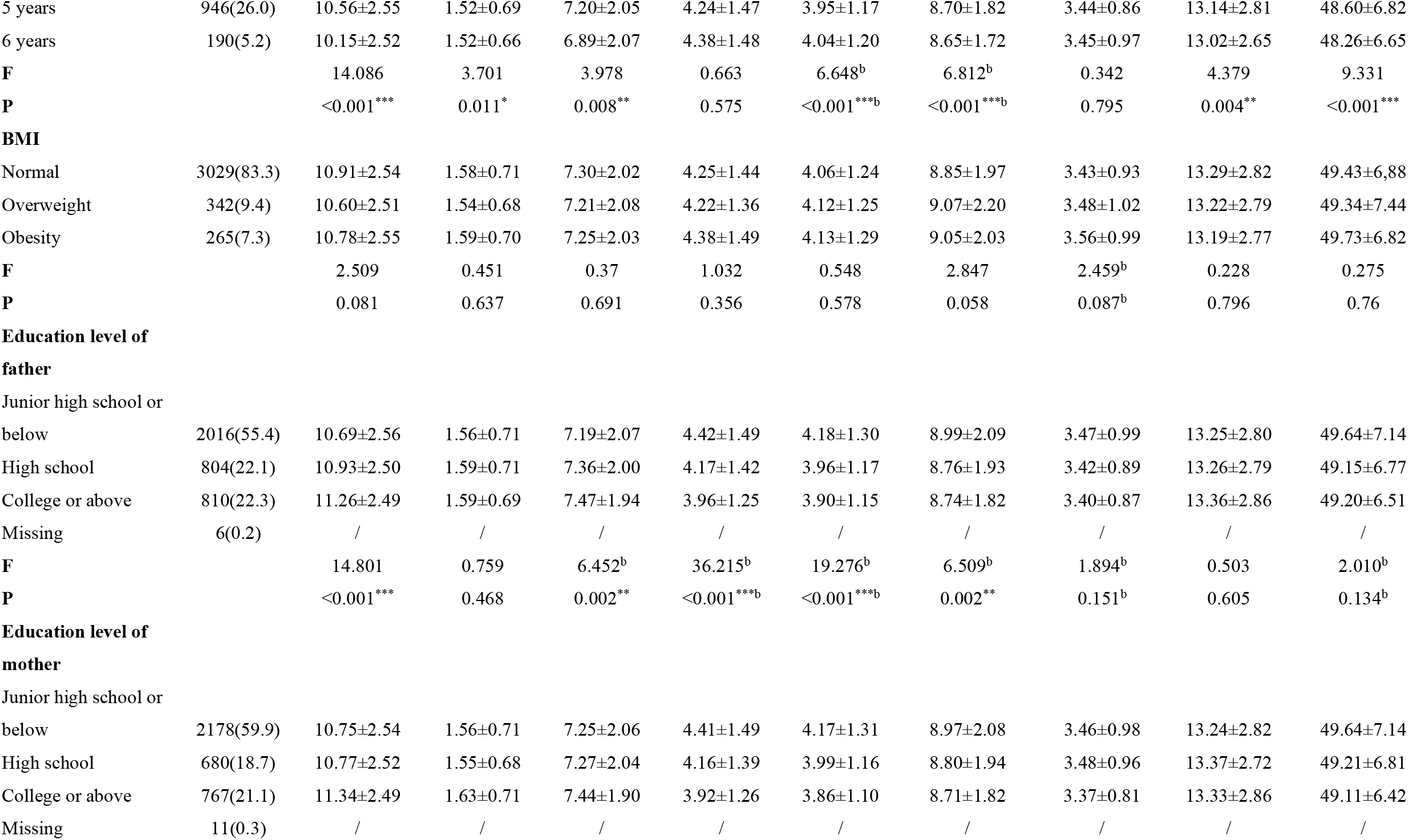
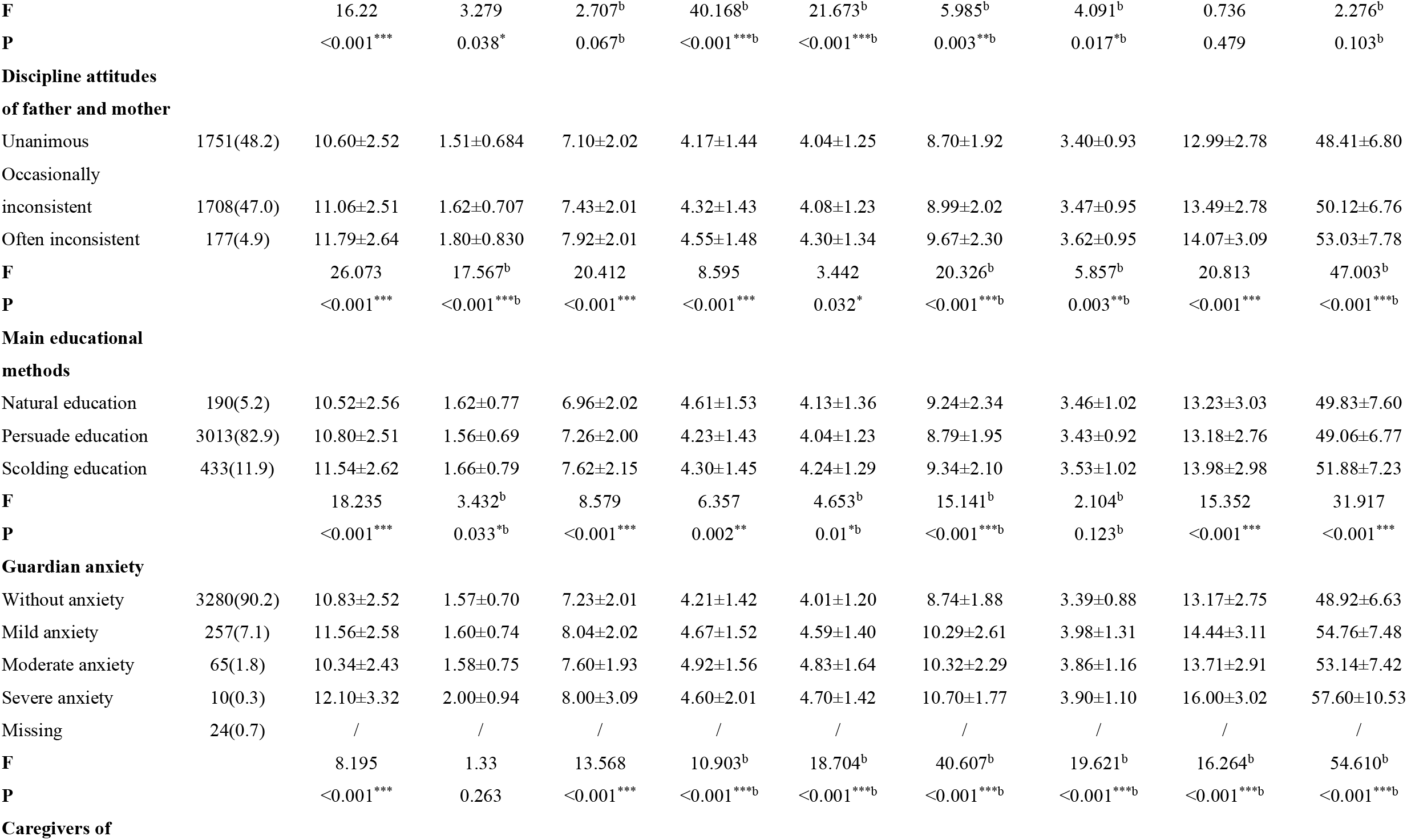
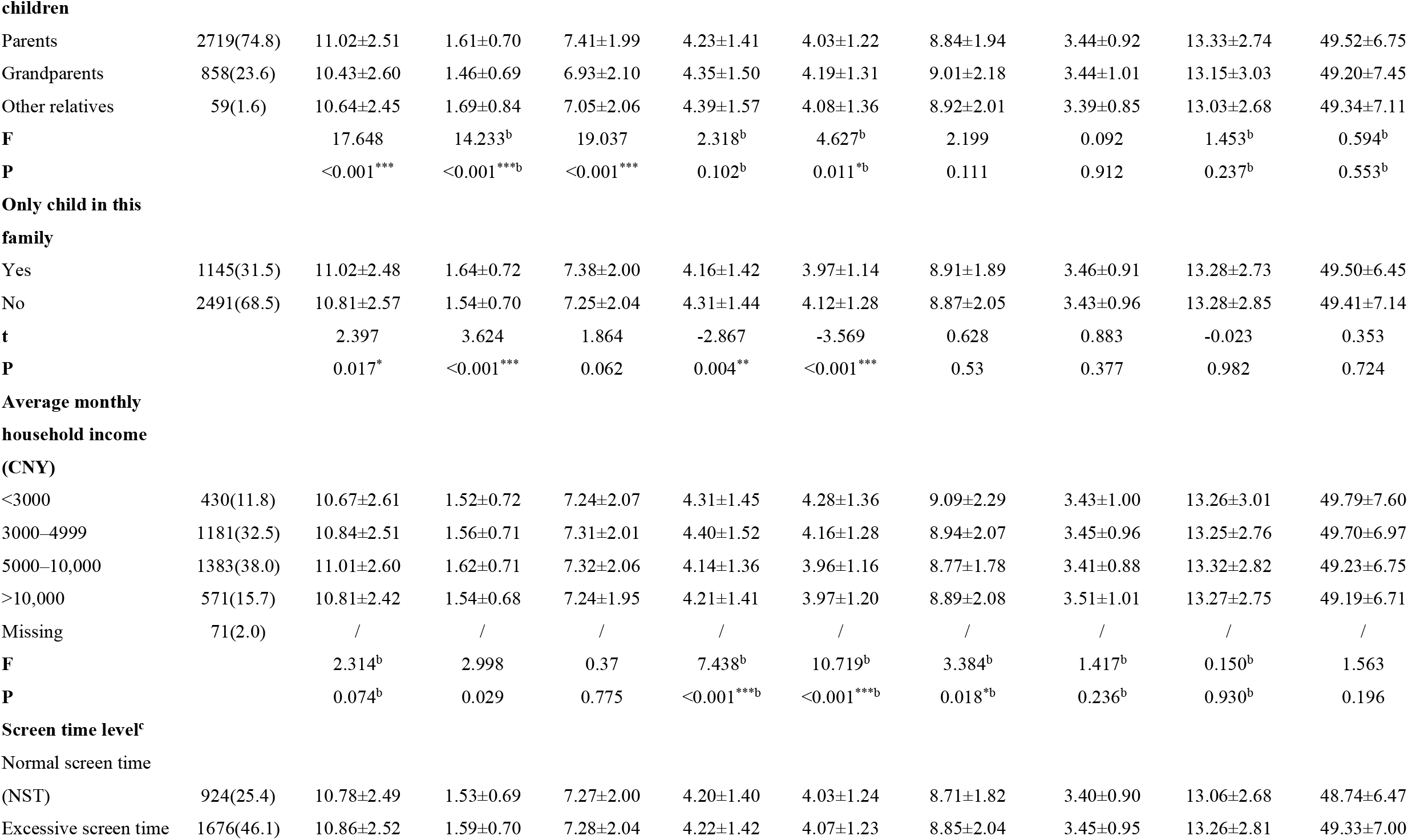
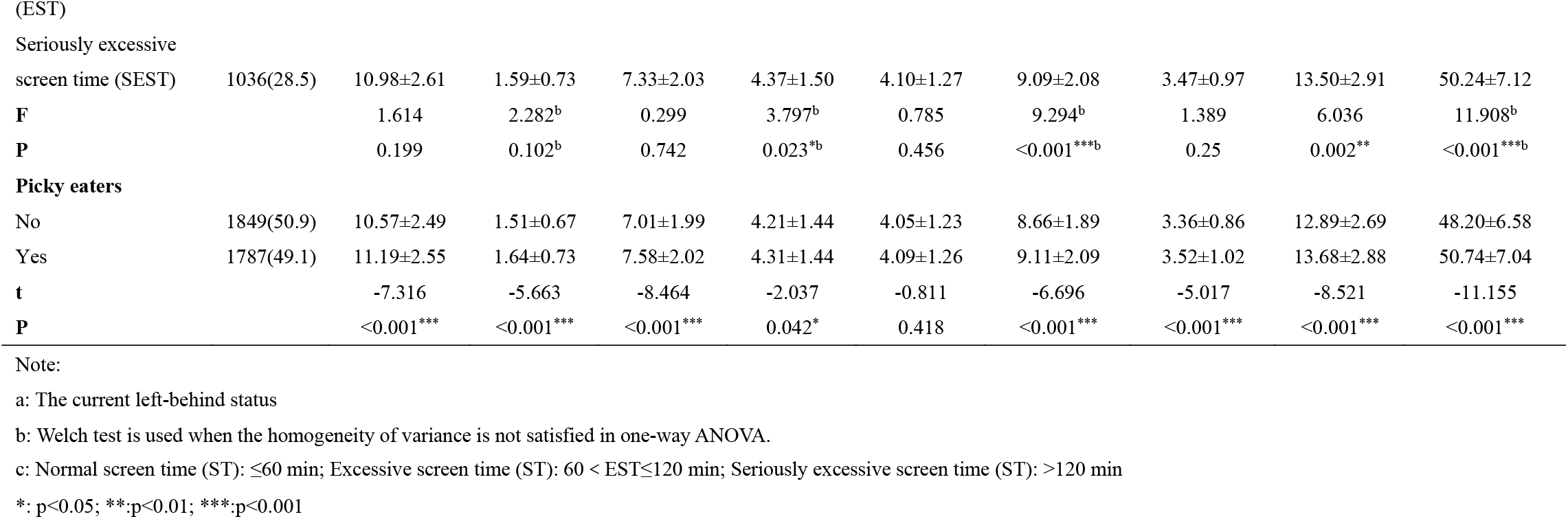
Comparison of CSHQ scores of preschool children with different characteristics.

As shown in Table 2, 3,214 subjects of our study scored above the CSHQ cutoff, indicating potential sleep disturbance. The prevalence of sleep disturbances among preschool children is 89.3%. As for the specific sleep disturbances based on each subscale, Bedtime Resistance (54.5%) and Sleep Anxiety (48.4%) are the most common type of sleep disturbances, which were followed by Daytime Sleepiness (21.5%), Sleep Duration (21.0%), Parasomnias (16.7%), Sleep Onset Delay (12.7%), Night Wakings (12.3%) and Sleep Disordered Breathing (10.5%).

**Table 2.**
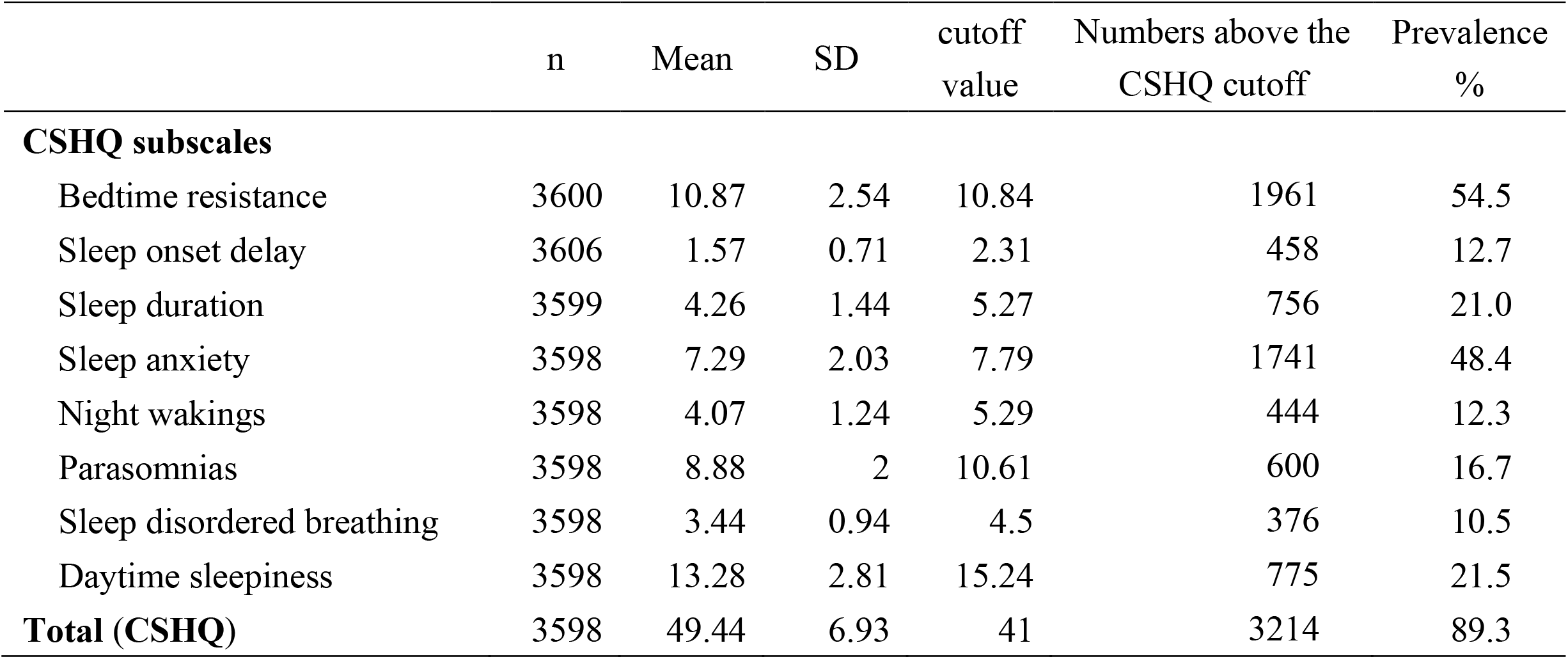
Sleep disturbances among preschool children

**Table 3.** Pearson correlation between subscales of CSHQ and SDQ

| **SDQ subscales** | **Total**<br>**(CSHQ)** | **Bedtime**<br>**Resistance** | **Sleep Onset**<br>**Delay** | **Sleep**<br>**Duration** | **Sleep**<br>**Anxiety** | **Night**<br>**Wakings** | **Parasomnias** | **Sleep**<br>**Disordered**<br>**Breathing** | **Daytime**<br>**Sleepiness** |
| --- | --- | --- | --- | --- | --- | --- | --- | --- | --- |
| Total (SDQ) | 0.365** | 0.168** | 0.091** | 0.174** | 0.168** | 0.17** | 0.284** | 0.222** | 0.216** |
| Hyperactivity/Inattention | 0.258** | 0.161** | 0.108** | 0.07** | 0.131** | 0.101** | 0.187** | 0.101** | 0.176** |
| Prosocial behavior | -0.176** | -0.066** | -0.063** | -0.135** | -0.034* | -0.063** | -0.145** | -0.098** | -0.113** |
| Conduct problems | 0.333** | 0.125** | 0.085** | 0.21** | 0.097** | 0.147** | 0.29** | 0.229** | 0.189** |
| Emotional symptoms | 0.357** | 0.157** | 0.069** | 0.172** | 0.169** | 0.172** | 0.265** | 0.238** | 0.215** |
| Peer problems | 0.216** | 0.057** | 0.043* | 0.192** | 0.068** | 0.104** | 0.184** | 0.145** | 0.113** |

\*. p<0.05; \*\*: p<0.01

### Correlation of subscales of the CSHQ and subscales of IMFeD as well as SDQ

As illustrated in Table 3, all the Pearson’s correlations between CSHQ and SDQ subscales were statistically significant (p<0.05 or <0.01). Besides, most of the correlations were in the positive direction except for the SDQ Prosocial subscale, which was inversely correlated. Nevertheless, most of correlations were low (r<0.3). Only the correlations of total CSHQ scores and SDQ scores (r=0.365), conduct problems (r=0.333) and emotional symptoms (r=0.357) were moderate. As for the Pearson’s correlations between CSHQ and IMFeD subscales, most of the Pearson’s correlations between CSHQ and IMFeD subscales were statistically significant (p<0.05 or <0.01) and all of them were inversely correlated. Only the correlations of total CSHQ scores and total IMFeD scores (r=-0.374) and fear of feeding (r=-0.305) were moderate (Table 4).

**Table 4.** Pearson correlation between subscales of CSHQ and IMFeD

|  | Total<br>(CSHQ) | Bedtime<br>Resistance | Sleep Onset<br>Delay | Sleep<br>Duration | Sleep<br>Anxiety | Night<br>Wakings | Parasomnias | Sleep Disordered<br>Breathing | Daytime<br>Sleepiness |
| --- | --- | --- | --- | --- | --- | --- | --- | --- | --- |
| Total (IMFeD) | -0.374** | -0.216** | -0.097** | -0.089** | -0.243** | -0.166** | -0.283** | -0.218** | -0.230** |
| Poor appetite | -0.279** | -0.195** | -0.088** | -0.012 | -0.208** | -0.113** | -0.211** | -0.156** | -0.174** |
| Food preference | -0.283** | -0.210** | -0.092** | -0.031 | -0.236** | -0.083** | -0.183** | -0.147** | -0.184** |
| Poor eating habit | -0.300** | -0.149** | -0.081** | -0.094** | -0.164** | -0.141** | -0.225** | -0.164** | -0.198** |
| Fear of feeding | -0.305** | -0.107** | -0.022 | -0.175** | -0.148** | -0.174** | -0.270** | -0.217** | -0.153** |
| Parental misperception | -0.276** | -0.163** | -0.083** | -0.061** | -0.182** | -0.126** | -0.197** | -0.139** | -0.181** |
| Organic disease | -0.280** | -0.076** | -0.006 | -0.149** | -0.097** | -0.204** | -0.289** | -0.274** | -0.102** |

\*. p<0.05; \*\*: p<0.01

### Hierarchical multiple regression analysis of sleep disturbances with different variables

Table 5 displays the results of hierarchical multiple regression analysis. In step 1, all sociodemographic variables account for about 5.3% (R^2^=0.053, p<0.01), which indicated that sociodemographic variables explained 5.3% of the variance of the CSHQ score. Among them, increasing of age (β=-0.693, p<0.01), discipline attitudes of father and mother (1=Unanimous, 2= occasionally inconsistent, 3=often inconsistent, β=1.822, p<0.01), main educational methods (1= natural education, 2= persuade education, 3= scolding education, β=1.683, p<0.01) and screen time level (0=NST, 1=EST, 2=SEST, β=0.673, p<0.01) predicted higher CSHQ scores significantly. In step 2, higher level of guardian anxiety also related to increasing CSHQ score (β=3.208, p<0.01) and this variable accounted for 3.1% of the variation of CSHQ score. In step 3 and 4, we found that all SDQ subscales and IMFeD subscales contributed to approximately 13.6% and 5.3% of the variation of CSHQ score respectively. In step 4, higher score of hyperactivity/Inattention (β=0.266, p<0.01), conduct problems (β=0.463, p<0.01), emotional symptoms (β=0.682, p<0.01) and peer problems (β=0.16, p<0.05) in SDQ suggested higher score of CSHQ, and lower score of prosocial behavior (β=-0.133, p<0.05) in SDQ as well as food preference (β=-0.151, p<0.01), fear of feeding (β=-0.262, p<0.01), parental misperception (β=-0.137, p<0.05) and organic disease (β=-0.728, p<0.01) in IMFeD were associated with higher score of CSHQ significantly.

**Table 5.** Associations between sociodemographic, anxious status of caregivers, emotional/behavioral problems as well as feeding difficulties and CSHQ based on hierarchical multiple regression analysis

| Variable | $\beta$ | | | |
| --- | --- | --- | --- | --- |
|  | step 1 | step 2 | step 3 | step 4 |
| **Sociodemographic factors** |  |  |  |  |
| Sex<sup>a</sup> | 0.198 | 0.144 | 0.279 | 0.219 |
| Left-behind status<sup>b</sup> | 0.547* | 0.383 | 0.072 | 0.094 |
| Birth weight<sup>c</sup> | -0.466 | -0.491 | -0.374 | -0.29 |
| Age (years) | -0.693** | -0.677** | -0.439** | -0.379** |
| BMI<sup>d</sup> | 0.042 | -0.046 | -0.16 | -0.003 |
| Education level of father<sup>e</sup> | -0.051 | 0.046 | 0.112 | 0.13 |
| Education level of mother<sup>f</sup> | -0.208 | -0.251 | -0.133 | -0.212 |
| Discipline attitudes of father and mother<sup>g</sup> | 1.822** | 1.567** | 0.819** | 0.756** |
| Main educational methods<sup>h</sup> | 1.683** | 1.518** | 0.829** | 0.724** |
| Caregivers of children<sup>i</sup> | -0.442 | -0.675* | -0.843** | -0.943** |
| Only child in this family<sup>j</sup> | -0.278 | -0.348 | -0.384 | -0.346 |
| Average monthly household income (CNY)<sup>k</sup> | -0.128 | -0.113 | 0.058 | 0.02 |
| Screen time level<sup>l</sup> | 0.673** | 0.622** | 0.277 | 0.195 |
| **Anxious status of caregivers** |  |  |  |  |
| Guardian anxiety<sup>m</sup> |  | 3.208** | 1.728** | 1.397** |
| **SDQ scales** |  |  |  |  |
| Hyperactivity/Inattention |  |  | 0.330** | 0.266** |
| Prosocial behavior |  |  | -0.183** | -0.133* |
| Conduct problems |  |  | 0.651** | 0.463** |
| Emotional symptoms |  |  | 0.840** | 0.682** |
| Peer problems |  |  | 0.208** | 0.16* |
| **IMFeD scales** |  |  |  |  |
| Poor appetite |  |  |  | -0.074 |
| Food preference |  |  |  | -0.151** |
| Poor eating habit |  |  |  | -0.05 |
| Fear of feeding |  |  |  | -0.262** |
| Parental misperception |  |  |  | -0.137* |
| Organic disease |  |  |  | -0.728** |
| **R<sup>2</sup>** | 0.053 | 0.086 | 0.221 | 0.274 |
| **F** | 14.355** | 22.214** | 49.529** | 49.847** |
| **R<sup>2</sup> change** | 0.053 | 0.032 | 0.136 | 0.053 |
| **F change** | 14.355** | 117.806** | 115.286** | 39.815** |

Note: \*\*p < 0.01; \*p < 0.05.
a: 1=male, 2=female; b: 0=non-LBC, 1=LBC; c: 1=underweight, 2=normal, 3=overweight; d: 0=normal, 1=overweight, 2=obesity; e: 1=junior high school or below, 2=high school, 3=college or above; f: 1=junior high school or below, 2=high school, 3=college or above; g: 1= unanimous, 2= occasionally inconsistent, 3= often inconsistent ; h: 1= natural education, 2= persuade education, 3= scolding education; i: 1=parents, 2=grandparents, 3=other relatives; j: 1=yes, 2=no; k: 1=<3000, 2=3000-4999, 3=5000-10,000, 4=>10,000; l: 0=normal screen time, 1= excessive screen time; 2= seriously excessive screen time; m: 0=without anxiety, 1=mild anxiety, 2=moderate anxiety, 3=severe anxiety.

## Discussion

In our study, we explored the prevalence of sleep disturbances and the associated factors comprehensively through several well-designed and validated instruments based on a larger sample in rural area of China. The associated factors related to sleep disturbance among preschool children living in rural areas may be different to that of urban preschool children on account of the difference in education and growth environment. Therefore, it is essential to carry out this exploratory research in order to supplement the results of previous studies [10].

The sleep disturbances of preschool children in rural area of China in our study was pretty high (89.3%) and the most common type was Bedtime Resistance and Sleep anxiety, which is consistent with previous studies [10]. Due to the cultural habits, co-sleeping is quite common in the Asian countries including China, Japan [9] and Korea [32], which partly contributed to the score of Bedtime Resistance and Sleep anxiety since these subscales shared some items related to co-sleeping (Needs parent in room to sleep; Afraid of sleeping alone). Besides, in some rural areas of China, many preschool children are usually nurtured by their grandparents or other relatives. They are more likely to take some comfort measures to satisfy the children so that they can sleep peacefully, and such inappropriate style may result in children prone to sleep problems. Such improper parenting methods will increase the risk of sleep problems, and the reduction of sleep quality will further cause a serious negative impact on the development of children’s physical and mental health. In our study, we find that the relationship of caregivers and children were associated with the CSHQ score of children in final model, compared with children nurtured by their parents, children whose caregivers were other relatives were tended to have less sleep disturbances. The potential reasons may as follows: 1. Other caregivers may pay less attention to children’s sleep problems than their parents and tend to give relatively lower scores when answering questions because of the lack of information; 2. Other caregivers didn’t want others to feel that they didn’t take care of the children well enough, which may bring a certain amount of information bias. 3. Children raised by other relatives may be more accustomed to sleep alone. The correlations between children’s age and total CSHQ score may result from developmental changes in sleep habits [9] due to the improvement of children’s knowledge and living ability. In terms of parental education style, our study found that inconsistent in discipline attitudes as well as scolding education were related to higher score of CSHQ compared to unanimous attitude and more lenient education style (persuade education). Previous studies have proven that inconsistent in child-rearing attitudes may increase the risk of emotional/behavioral problems in children, which are related to sleep disturbances [10, 33]. Furthermore, parents should maintain the consistency in their educational theory in order to prevent potential confusion and anxiety in their children which may related to their sleep quality; besides, instead of scolding or beating, education methods should mainly base on persuasion and setting an example in case of adverse effects in children’s physical and mental health which are related to their sleep quality as well.

Our survey found that the anxious level of caregivers was positively related to the children’s CSHQ score, the higher a caregivers’ anxiety level, the more sleep problems their child had. Previous studies have shown that the association between parental anxiety and sleep problems of children may be bidirectional [1, 4, 34, 35]. Children are likely to be exposed to parental mood disturbances which could create an environment of uncertainty and insecurity that may increase the risk of sleep problems [1, 36]. Furthermore, existing of sleep problems in children may make their parents feel more stressful and are more likely to feel distressed [37] which could generate or even aggravate their anxiety. Our study indicated the importance of psychological intervention of caregivers on improving their children’s sleep quality.

In our study, we found significant correlations between SDQ subscales (Emotional Symptoms, Conduct Problems, Hyperactivity /Inattention, Peer Relationship Problems and Prosocial Behavior) and sleep disturbances among preschool children. Compared with previous studies in school-aged children [22] and urban preschool children [10], we also found significant correlations between prosocial behavior and CSHQ score. Results from our study indicated that the characteristics of sleep disturbances in preschool children in rural areas may differ from that of urban preschoolers and the potential association between prosocial behaviors and sleep disturbances in preschool children should be paid more attention to in studying the emotional and behavioral problems of children.

In addition, our survey suggested that feeding difficulties may influence the children’s sleep quality. Food preference, fear of feeding, parental misperception and organic disease were related to sleep disturbances amongst children. In rural areas of China, the eating perception of caregivers may have some distinct characteristics: due to relatively lower level of education, caregivers may believe that overweight children will be more healthy and taller in the future [31, 38], which could make them overfeed and care excessively about the children’s eating and feeding [31]; this tendency could both lead to feeding difficulties in children and wrong perspective (thinking their children did not eat enough) of caregivers. Previous study demonstrated that shorter nocturnal sleep duration was related to overeating in the absence of hunger [39], and children with unhealthy diet including eating outside the home, eating fast foods, eating in front of the TV or eating alone are more likely to have shorter sleep and poorer sleep quality [40]. Healthy eating habits are essential to the physical and mental development of preschool children which could influence the sleep quality of children. Our study emphasized the importance of healthy eating habits for improving the sleep quality of preschoolers potentially. Building healthy eating habits and establishing a correct concept of feeding for caregivers especially in rural areas are beneficial to the physical and mental health of preschool children which could further improve their sleep quality.

However, our study has also some limitations. Firstly, limited by the cross-sectional study design, our study result cannot ensure the causal relationship between sleep disturbances of preschool children and associated factors discovered in our study, future well-designed cohort studies are needed to verify the results of our research. Secondly, our study sites were selected from rural areas of Anhui province in China, whose generalizability to other populations located in other provinces in China or abroad was limited. Thirdly, based on the number of variables and scales included in our study, our study aimed to explore the potential factors related to sleep disturbances among preschool children, which was limited by specific and deep analysis on certain factors. Finally, limited by the education level of caregivers in our study, parents or other caregivers may not give the information of their children precisely, which may cause certain amount of information bias.

In conclusion, the prevalence of sleep disturbances among preschool children in rural areas of China was quite high and the potential risk factors were multidimensional including children-related factors (age, emotional and behavioral problems and feeding difficulties, etc.) and caregivers’ factors (discipline attitudes, main educational methods, main caregivers, anxiety status of caregivers, etc.). Future well-designed and large-sampled studies are needed to explore the specific risk factors and explore targeted interventions both for children and their caregivers.

## Data Availability

The raw data required to reproduce these findings cannot be shared at this time as the data also forms part of an ongoing study.

## Acknowledgements

We appreciate the authors listed in this paper for their contributions to this study, including study design, site investigation, data collation and analysis, and paper writing; in addition, we also appreciate the local Center for Disease Control and Prevention and all kindergartens for their great cooperation. Finally, Our gratitude to the Professor Yehuan Sun for his constructive comments and final revision to this paper.

